# The Role of PITPNC1 in Lung Adenocarcinoma: Differential Expression, Immune Infiltration, and Prognostic Significance

**DOI:** 10.1101/2024.08.18.24312183

**Authors:** Chao Li, Junsong Chen, Ganggang Zhang, Fang Guo, Xin Zhang

**Affiliations:** Department of Pulmonary Medicine, Wuhu No. 1 People’s Hospital, Wuhu, PR China; Key Laboratory of Systems Biomedicine (Ministry of Education), Shanghai Center for Systems Biomedicine, Shanghai Jiao Tong University, Shanghai, PR China

**Keywords:** Lung adenocarcinoma(LUAD), PITPNC1, bioinformatics analysis, metastasis

## Abstract

Lung adenocarcinoma (LUAD) is one of the most prevalent and deadly forms of lung cancer, necessitating the identification of novel biomarkers for diagnosis and prognosis. This study aims to explore the differential expression, diagnostic potential, underlying mechanisms, and clinical significance of PITPNC1 (phosphatidylinositol transfer protein, cytoplasmic 1) in LUAD.We utilized data from The Cancer Genome Atlas (TCGA) and Gene Expression Omnibus (GEO) databases, comprising 530 LUAD samples and 59 control samples from TCGA-LUAD, as well as GSE10072 and GSE75037 datasets with a total of 224 samples. Data preprocessing included normalization to Fragments Per Kilobase of transcript per Million mapped reads (FPKM) format and batch effect correction using the R package sva. Differential gene expression analysis was performed using DESeq2 for TCGA-LUAD and limma for GEO datasets. Receiver Operating Characteristic (ROC) curve analysis was conducted to assess the diagnostic efficacy of PITPNC1.Our results revealed that PITPNC1 is significantly overexpressed in LUAD samples compared to controls (p < 0.001 in TCGA-LUAD; p < 0.01 in GEO). However, ROC curve analysis indicated moderate diagnostic accuracy with Area Under Curve (AUC) values between 0.5 and 0.7. Differential expression analysis identified 3838 genes associated with PITPNC1 expression, which were further subjected to Gene Ontology (GO) and Kyoto Encyclopedia of Genes and Genomes (KEGG) enrichment analyses. These genes were enriched in pathways related to external stimulus response, hormone level regulation, nitrogen metabolism, and neuroactive ligand-receptor interaction.Gene Set Enrichment Analysis (GSEA) highlighted significant enrichment in IL12 signaling pathway, Notch signaling pathway, MAPK6/MAPK4 signaling pathway, and Hedgehog On State pathway. Immune infiltration analysis using single-sample Gene Set Enrichment Analysis (ssGSEA) showed significant differences in five immune cell types between high and low PITPNC1 expression groups. Cox regression analysis indicated that PITPNC1 expression along with clinical stages are significant predictors of overall survival in LUAD patients.In conclusion, our comprehensive bioinformatics analysis underscores the potential role of PITPNC1 as a biomarker for LUAD diagnosis and prognosis.

## 1. INTRODUCTION

Lung adenocarcinoma (LUAD) represents the predominant subtype of non-small cell lung cancer (NSCLC) and continues to be a significant contributor to cancer-related deaths globally.In 2020, lung cancer accounted for approximately 2.2 million new cases and 1.8 million deaths worldwide, with adenocarcinoma being the predominant histological type[1]. Despite advancements in diagnostic techniques and therapeutic strategies, the prognosis of LUAD patients remains poor, advanced NSCLC has a 5-year survival rate less than 20% [2]. Current treatments, including surgery, chemotherapy, radiotherapy, and targeted therapies, often face limitations such as drug resistance and adverse effects, underscoring the need for novel diagnostic and therapeutic markers[3].

PITPNC1 (phosphatidylinositol transfer protein, cytoplasmic 1) has been identified as a potential oncogene in various cancers, including gastric cancer and colorectal cancer and melanoma cells, Halberg et al. identified PITPNC1 as a gene amplified in a significant portion of human breast cancer and observed its overexpression across multiple types of cancer [4], where it plays a role in tumor progression and metastasis[5]. Its involvement in LUAD, however, has not been extensively studied. In gastric cancer, PITPNC1 promotes cell invasion and proliferation by activating the PI3K/AKT signaling pathway[6]. These findings indicate that PITPNC1 could serve as a valuable biomarker and therapeutic target in LUAD.

This study aims to explore the expression, diagnostic potential, and underlying mechanisms of PITPNC1 in LUAD. We examined the varying expression levels of PITPNC1 in both LUAD and normal lung tissues by utilizing information from the TCGA and GEO databases. Furthermore, we assessed the diagnostic performance of PITPNC1 through ROC curve analysis and examined its association with immune infiltration and clinical outcomes. Functional enrichment analyses, including GO, KEGG, and GSEA, were performed to elucidate the biological processes and pathways involving PITPNC1. Additionally, we constructed protein-protein interaction (PPI) networks and regulatory networks to identify key interacting partners and regulatory elements associated with PITPNC1. Finally, we evaluated the prognostic value of PITPNC1 using survival analysis and developed a nomogram to predict overall survival in LUAD patients.

In summary, our comprehensive bioinformatics analysis revealed that PITPNC1 is significantly overexpressed in LUAD and may serve as a promising diagnostic and prognostic biomarker. The findings also suggest that PITPNC1 is involved in critical oncogenic pathways and immune regulation, offering potential therapeutic targets for future interventions and delivering new perspectives on the molecular mechanisms that drive the progression of LUAD.

## 2. MATERIALS AND METHODS

### 2.1 Data Download

We downloaded Lung Adenocarcinoma from The Cancer Genome Atlas (TCGA) via the R package TCGAbiolinks[7] (Version 2.30.0). The TCGA-LUAD dataset was analyzed as a test set. After excluding data samples without prognostic information, 530 LUAD samples with prognostic information and 59 Control samples were obtained in Counts format sequencing data. At the same time, it was standardized to FPKM (Fragments Per Kilobaseper Million) format, and the corresponding clinical data were obtained through UCSC Xena database[8], the specific information is shown in Table 1.

**Table 1.** Baseline Table with LUAD Patients Characteristics.

| Characteristics | overall |
| --- | --- |
| Age, median (IQR) | 66 (59, 72) |
| Gender, n (%) |  |
| MALE | 247 (46.6%) |
| female | 283 (53.4 percent) |
| NStage, n (%) |  |
| N0 | 345 (65.2%) |
| N1 | 96 (18.1%) |
| N2&3 | 73 (13.8 percent) |
| NX | 15 (2.8%) |
| TStage, n (%) |  |
| T1 | 176 (33.2%) |
| T2 | 285 (53.8%) |
| T3&4 | 66 (12.5%) |
| TX | 3 (0.6%) |
| Stage, n (%) |  |
| I | 292 (55.9%) |
| III&IV | 107 (20.5%) |
| II | 123 (23.6%) |

LUAD, Lung Adenocarcinomas.

Through the R package GEOquery[9] (Version 2.70.0) from GEO database[10] to download LUAD GSE10072[11], GSE75037[12] dataset. The samples of datasets GSE10072 and GSE75037 were all from Homo sapiens, and the tissue source was Lung; the specific information is shown in Table 2. Among them, the chip platform of dataset GSE10072 was GPL96, which included 58 LUAD samples and 49 Control samples. The chip platform of dataset GSE75037 was GPL6884, which included 83 LUAD samples and 83 Control samples. All samples were included in this study.

**Table 2.** GEO Microarray Chip Information.

|  | GSE10072 | GSE75037 |
| --- | --- | --- |
| Platform | GPL96 | GPL6884 |
| Species | Homo sapiens | Homo sapiens |
| Tissue | Lung | Lung |
| Samples in LUAD group | 58 | 83 |
| Samples in Control group | 49 | 83 |
| Reference | PMID: 18297132 | PMID: 27354471 |

GEO, Gene Expression Omnibus; LUAD, Lung Adenocarcinoma.

The R package sva[13] (Version 3.50.0) was used to debatching the Datasets GSE10072 and GSE75037 to obtain the Combined GEO datasets. Among them, the Combined Datasets included 141 cases and 132 controls. Finally, the Combined Datasets were standardized by R package limma[14] (Version 3.58.1), and the annotation probes were standardized and normalized. Principal component analysis(PCA) was performed on the matrix of equations before and after eliminating batch effects to verify the effect of removing batch effect [15]. PCA is a data dimensionality reduction method that extracts data feature vectors (components) from high-dimensional data. To transform the data into low-dimensional data and display these features in 2D or 3D graphs.

### 2.2 Lung adenocarcinoma-associated differentially expressed genes

Based on the sample classification of the lung adenocarcinoma dataset (TCGA-LUAD), the specimens were categorized into the LUAD group and the Control group. The differential analysis of genes in the LUAD group compared to the Control group was carried out using the R package DESeq2 (Version 1.42.0)[16]. The threshold for Differentially Expressed Genes (DEGs) is established with logFC > 0 and adjusted P < 0.01 for genes that are up-regulated, while logFC < 0 and adjusted P < 0.01 indicate down-regulated genes. The method employed for p-value correction was the Benjamini-Hochberg (BH) approach. The outcomes of the differential analysis were visualized utilizing the R package ggplot2 (Version 3.4.4) to create volcano plots.

Based on the classification of the Combined Datasets, the samples were separated into the LUAD group and the Control group. The differential gene analysis between the LUAD group and the control group was conducted using the limma package (Version 3.58.1) in R. The threshold for DEGs is defined as logFC > 0 and adjusted P < 0.01 for up-regulated genes, while down-regulated genes are characterized by logFC < 0 and adjusted P < 0.01. p value correction method was Benjamini-Hochberg (BH). The results of the difference analysis were plotted using the R package ggplot2 (Version 3.4.4) for volcano plots.

To identify differentially expressed genes associated with LUAD, All up-regulated DEGs with logFC > 0 and adj.p < 0.01 obtained by differential analysis in the lung adenocarcinoma dataset (TCGA-LUAD) and the GEO Datasets (Combined Datasets) were interposed and Venn diagram was drawn. Then, differential analysis of theTCGA-LUAD and combined GEO datasets yielded all up-regulated DEGs with logFC > 0 and adj.p < 0.01, which were then interspersed, allowing for the creation of a Venn diagram. Genes that are differentially expressed concerning LUAD were identified by merging the intersecting genes, and a heatmap was generated using the R package pheatmap (Version 1.0.12).

### 2.3 Differential expression verification and ROC curve analysis of PITPNC1

In order to further study the expression differences of PITPNC1 in the LUAD group and the Control group of the TCGA-LUAD and the Combined GEO Datasets. The statistical significance of differences was estimated using independent Student’s t test according to PITPNC1 expression levels, and comparison plots between groups were constructed. The ROC curve of PITPNC1 was plotted on the TCGA-LUAD and GEO combined data sets using the R software package pROC (version 1.18.5), and the area under the ROC curve (AUC) was calculated and used to estimate expression levels. I calculated the AUC, PITPNC1, diagnostic role in the development of LUAD. The area under the receiver operating characteristic curve is usually between 0.5 and 1. AUC values close to 1 indicate better diagnostic performance.An AUC between 0.5 and 0.7 indicates low accuracy, an AUC between 0.7 and 0.9 indicates moderate accuracy, and an AUC above 0.9 indicates high accuracy.

### 2.4 Differential analysis and co-expressed genes of PITPNC1 high and low expression groups

To explore the differentially expressed genes in LUAD between the PITPNC1 high expression group and the low expression group, as well as the underlying mechanism and related biological characteristics and pathways. Based on the median Expression of PITPNC1 in the LUAD samples of the TCGA-LUAD, we divided the LUAD samples into high expression group and low expression group. Difference analysis was performed using the R packages DESeq2 (version 1.42.0) and |logFC|. Match > 1.5 and aj. DEG threshold was set at P < 0.01. Contains logFC > 1.5 and adj. Genes with P < 0.01 are differentially up-regulated genes (up-regulated genes). logFC <-1.5 and adj.p <0.01 are downregulated genes. The results of the differential analysis were displayed by plotting volcano plots using the R package ggplot2 (version 3.4.4).Subsequently, heatmaps of expression values of differentially expressed genes were created using the R package pheatmap (version 1.0.12).

Next, in order to obtain the Co-expressed Genes of PITPNC1, we performed pairwise correlation analysis between PITPNC1 and other DEGs in the LUAD samples of the TCGA-LUAD dataset. The Co-expressed genes were identified as the top 20 genes correlated with PITPNC1, and the correlation heat map along with the co-expression heat map was created utilizing the R package ggplot2 (Version 3.4.4).

### 2.5 Gene ontology (GO) and pathway (KEGG) enrichment analysis

Gene Ontology (GO) analysis[17] serves as a prevalent approach for extensive functional enrichment investigations, which encompass Biological Process (BP), Cellular Component (CC), and Molecular Function (MF). The Kyoto Encyclopedia of Genes and Genomes (KEGG)[18] is a commonly utilized database that catalogs details regarding genomes, biological pathways, diseases, and drugs. In our study, we employed the R package clusterProfiler[19] (Version 4.10.0) to perform Gene ontology and pathway enrichment analysis of the DEGs from the differential analysis of high and low expression groups. p < 0.05 and FDR value (q value) < 0.25 were considered to be statistically significant, with the Benjamini-Hochberg (BH) method applied for p-value correction.

### 2.6 Gene Set Enrichment Analysis (GSEA)

GSEA[20] is utilized to assess the pattern of gene distribution within a predefined gene set, which is organized in a gene table based on its correlation with the phenotype, thereby identifying their impact on the phenotype. In this study, the genes in the LUAD samples of the TCGA-LUAD were ranked according to logFC values, and then, The R package clusterProfiler (Version 4.10.0) was used to perform GSEA on all genes in LUAD samples.The parameters utilized in GSEA were defined as follows: the seed value set was 2020, with a minimum of 10 genes and a maximum of 500 genes in each gene set. Access to the c2 gene sets was obtained from the Molecular Signatures Database (MSigDB). Specifically, the version used was Cp. All. V2022.1. Hs. Symbols. The GMT file containing [all Canonical Pathways](3050) was employed. The criteria for screening in GSEA included an adjusted p < 0.05 and FDR value (q value) < 0.25,with the p-value adjustment performed using the Benjamini-Hochberg (BH) method.

### 2.7 Protein-protein Interaction (PPI) Network

PPI Network, frequently referred to as the protein-protein interaction network, is made up of proteins that engage with one another, playing roles in biological signaling, gene expression regulation, and various life processes such as metabolism of energy and substances, as well as cell cycle regulation. Conducting a systematic analysis of protein interactions within biological systems holds significant importance for grasping how proteins function within these systems, elucidating the mechanisms behind biological signaling and the metabolism of energy and substances, especially under specific physiological conditions like diseases, and uncovering the functional relationships among proteins. The GeneMANIA database[21] serves to hypothesize gene functions, conduct gene list analyses, and prioritize genes for further functional investigations. When provided with a list of query genes, GeneMANIA identifies functionally analogous genes utilizing an extensive range of genomics and proteomics data. In this approach, each functional genomic dataset is assigned a weight that corresponds to the expected value of the query. Additionally, GeneMANIA can predict gene functions; given a query gene, it identifies other genes likely to have shared functions based on their interactions. A PPI Network was developed by forecasting the functionally similar genes of PITPNC1 via the GeneMANIA online platform.

### 2.8 Construction of regulatory network

Transcription factors (TFS) regulate gene expression by interacting with PITPNC1 at post-transcriptional steps. We searched for transcription factors (TF) in the ChIPBase database [22], analyzed the regulation of transcription factors (TF), and transcribed PITPNC1-regulated genes into mRNA. -TF. Cytoscape [23] constructs networks through visualization software. In addition, miRNA plays an important regulatory role in biological development and evolution. It can be used to regulate many target genes,and the same target gene can also be regulated by multiple miRNAs. In order to analyze the relationship between the gene PITPNC1 and microRNAs, through a StarBase v3.0 database.The mRNA-miRNA regulatory network was visualized using Cytoscape software

### 2.9 Immune infiltration analysis

Single-Sample Gene-Set Enrichment Analysis (ssGSEA) [25] is a methodology utilized to quantify the relative abundance of various immune cell infiltrates within individual samples. Initially, distinct types of infiltrating immune cells were categorized, including Activated CD8 T cells, Activated dendritic cells, Gamma delta T cells, Natural killer cells, and various subtypes of human immune cells such as Regulatory T cells. The enrichment scores derived from the ssGSEA analysis were subsequently employed to indicate the relative abundance of immune cell infiltration across each sample, thereby generating an immune cell infiltration matrix for LUAD samples sourced from the TCGA-LUAD.Following this, the R package ggplot2 (Version 3.4.4) was utilized to create comparative visualizations that illustrate the expression disparities of immune cells between the high expression and low expression groups within the LUAD samples. Immune cells exhibiting significant differences between these two groups were identified for further analysis, and the correlation among immune cells was assessed using the Spearman algorithm. To represent the correlation analysis results visually, the R package pheatmap (Version 1.0.12) was employed to generate a correlation heatmap. Additionally, the Spearman algorithm was applied to evaluate the correlation between PITPNC1 and the various immune cells, with the correlation bubble plot created using the R package ggplot2 (Version 3.4.4) to depict the results of this correlation analysis.

### 2.10 Construction of clinical prognostic model and prognostic analysis of lung adenocarcinoma

The time-dependent Receiver Operating Characteristic Curve (ROC) [26]serves as a graphical tool for model selection, enabling the identification of the optimal model, the exclusion of inferior models, or the establishment of the best threshold within a given model. The R package survivalROC (Version 1.0.3.1) was employed to generate the time-dependent ROC Curve and to compute the Area Under the Curve (AUC) for PITPNC1 expression in relation to overall survival (OS) among LUAD samples. This analysis aimed to predict the survival outcomes at 1, 2, and 3 years for LUAD samples from the TCGA-LUAD dataset. Generally, the AUC values of the ROC curve range from 0.5 to 1, with values closer to 1 indicating superior diagnostic performance. An AUC greater than 0.5 suggests a tendency for the expression of the molecule to promote the occurrence of the event; an AUC between 0.5 and 0.7 indicates low accuracy, between 0.7 and 0.9 signifies moderate accuracy, and above 0.9 reflects high accuracy.

To investigate the differences in overall survival (OS) between the high expression and low expression groups of LUAD samples within TCGA-LUAD, a Kaplan-Meier (KM) curve[27] analysis was conducted utilizing the R package survival (Version 3.5-7). The KM curve was generated based on the expression levels of the PITPNC1 gene in LUAD samples.

The findings from both univariate and multivariate Cox regression analyses were illustrated using a Forest Plot, which highlighted the expression levels of the PITPNC1 gene alongside relevant clinical information from the LUAD samples analyzed in these regression models. A Nomogram, which is a graphical representation employing a series of disjoint line segments to depict the functional relationships among multiple independent variables within a rectangular coordinate system, was constructed using the R package "rms" (Version 6.7-1). This Nomogram elucidated the relationship between PITPNC1 expression levels and clinical data in LUAD samples incorporated in the multivariate Cox regression analysis.

Additionally, the R package "ggplot2" (Version 3.4.4) was employed to create a risk factor map based on the expression levels of PITPNC1 in LUAD samples. To assess the predictive performance of the model concerning actual outcomes, a Calibration Curve was generated. This curve plotted the correspondence between the actual probabilities and the probabilities predicted by the model under varying conditions. Calibration analysis was performed to evaluate the accuracy and discriminative ability of the prognostic risk model grounded in PITPNC1 gene expression.

### 2.11 Statistical analysis

The data processing and analytical procedures described in this study were conducted utilizing R software (Version 4.3.0). When comparing continuous variables across two groups, the statistical significance of normally distributed variables was assessed using the independent Student’s T-Test, unless stated otherwise. For variables that did not conform to a normal distribution, the Mann-Whitney U Test (also known as the Wilcoxon Rank Sum Test) was employed to evaluate differences. The Kruskal-Wallis test was applied for the comparison of three or more groups. To determine the correlation coefficient among various molecules, Spearman correlation analysis was utilized. Unless indicated otherwise, all statistical p-values were two-sided, with a threshold of p < 0.05 denoting statistical significance.

## 3. RESULTS

### 3.1 Technology Roadmap

### 3.2 Merging of lung adenocarcinoma datasets

Firstly, the R package sva was used to perform batch effect removal on LUAD Datasets GSE10072 and GSE75037 to obtain Combined GEO datasets. Subsequently, the distribution boxplots (Fig 2A-B) were used to compare the expression values of the datasets prior to and following batch effect. Secondly, PCA plot (Fig 2C-D) was used to compare the distribution of low-dimensional features before and after batch effect removal. The results of distribution boxplot and PCA plot indicated that the batch effect of the LUAD dataset was basically eliminated after removing the batch.

**Fig. 1.**
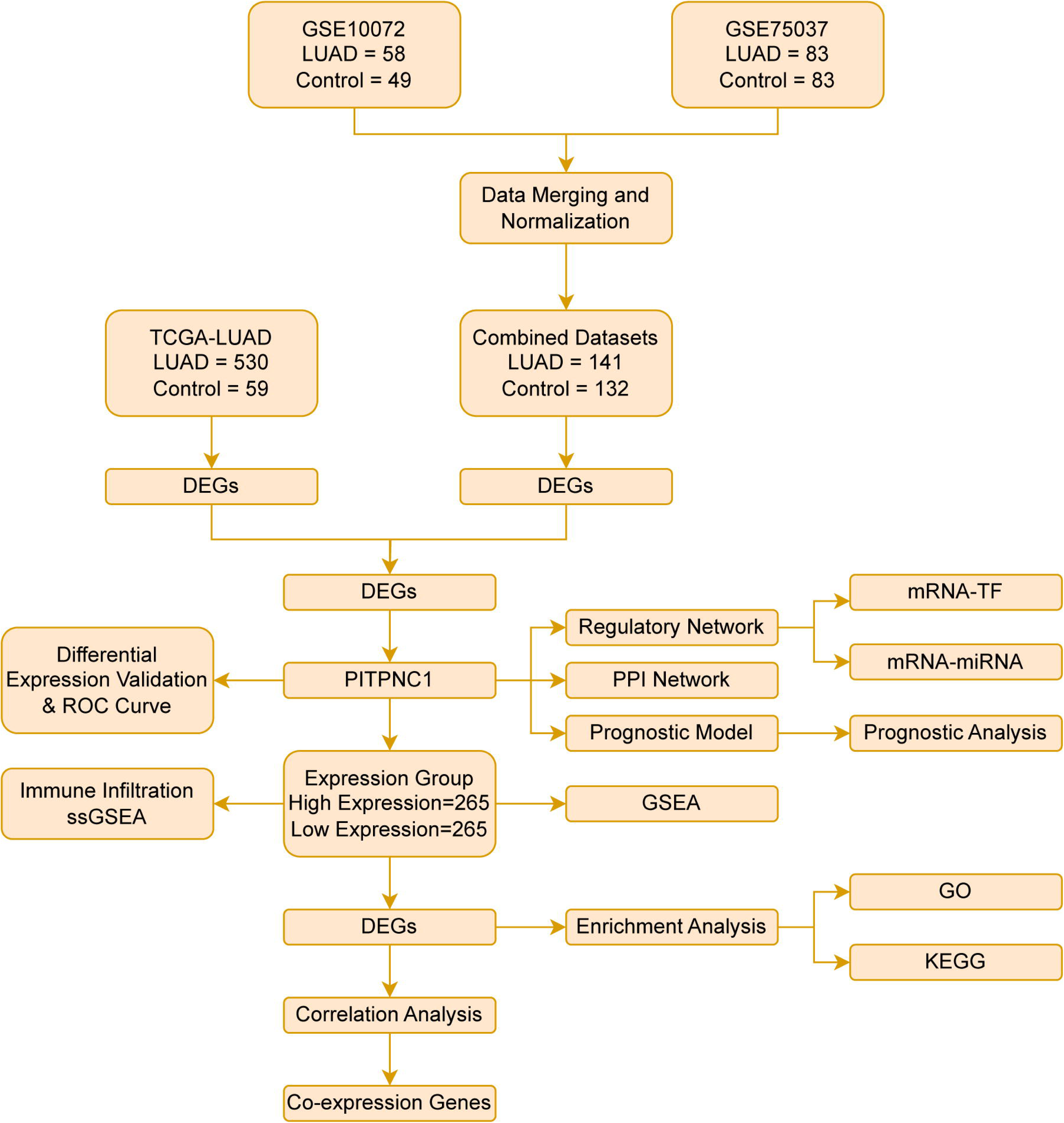
Flow Chart for the Comprehensive Analysis of PITPNC1. TCGA, The Cancer Genome Atlas; LUAD, Lung Adenocarcinoma; DEGs, Differentially Expressed Genes; PPI, Protein-protein Interaction; ROC, Receiver Operating Characteristic; TF, Transcription Factor; GSEA, Gene Set Enrichment Analysis; ssGSEA, Single-Sample Gene-Set Enrichment Analysis; GO, Gene Ontology; KEGG, Kyoto Encyclopedia of Genes and Genomes.

**Fig. 2.**
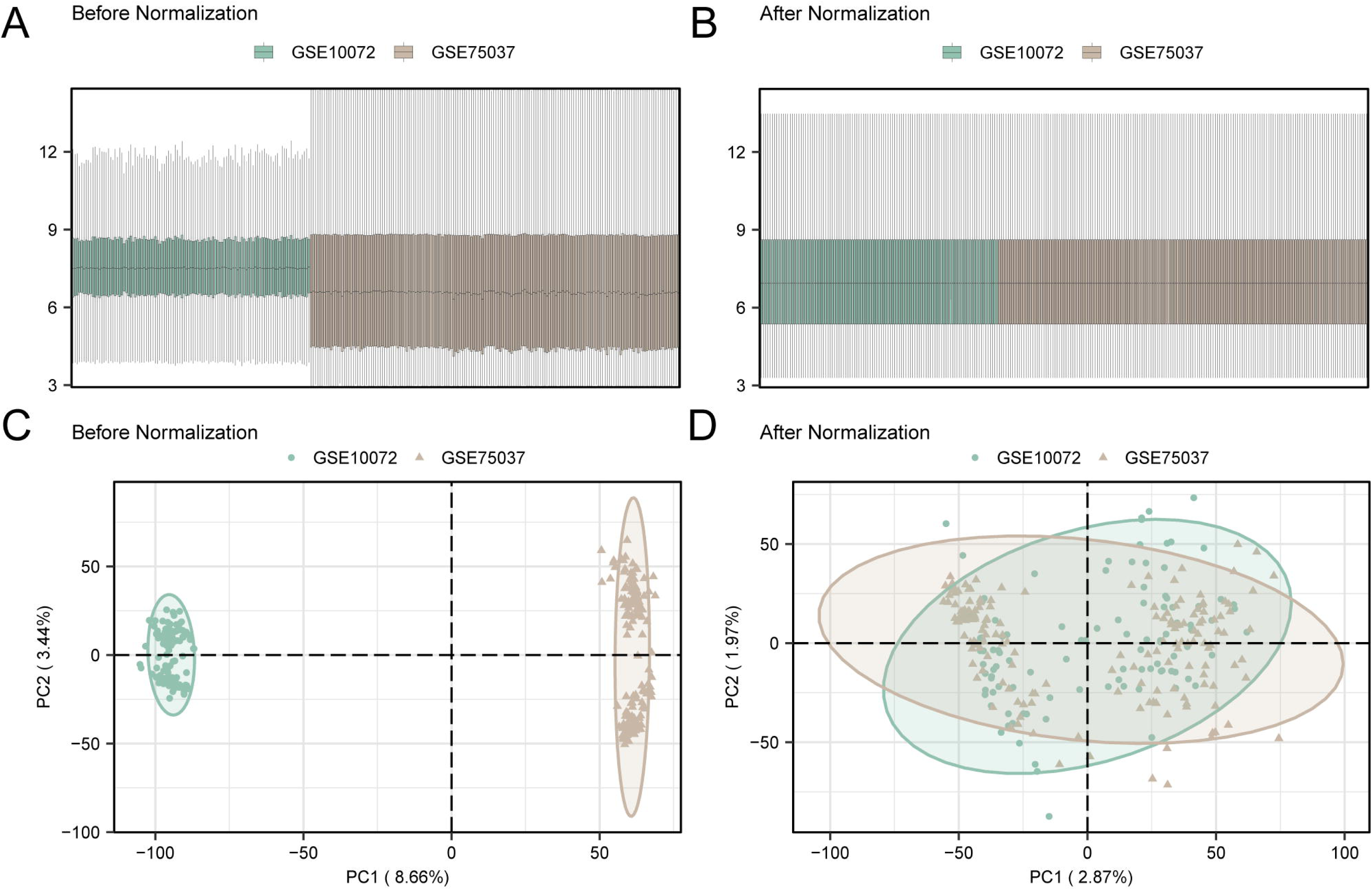
Batch Effects Removal of GSE10072 and GSE75037. A. Box plot of Combined GEO Datasets distribution before batch removal. B. Post-batch integrated GEO Datasets (Combined Datasets) distribution boxplots. C. PCA plot of integrated GEO Datasets (Combined Datasets) before debatching. D. PCA plot of integrated GEO Datasets (Combined Datasets) after debatching. PCA, Principal Component Analysis; LUAD, Lung Adenocarcinoma. The lung adenocarcinoma (LUAD) dataset GSE10072 is green, and the lung adenocarcinoma (LUAD) dataset GSE75037 is brown.

### 3.3 Lung adenocarcinoma-related differentially expressed genes

The TCGA-LUAD data were divided into LUAD group and Control group. In order to analyze the difference of gene expression values between the LUAD group and the Control group in the TCGA-LUAD, the R package DESeq2 was used for differential analysis of the TCGA-LUAD to obtain the differentially expressed genes in the two groups of data, and the results are as follows: TCGA -LUAD a total of 23873 data sets meet | logFC | > 0 and adj. P < 0.01 threshold of DEGs; Under the threshold, raised expressed genes (logFC > 0 and adj. P < 0.01), a total of 17336 cut expressed genes logFC < 0 and adj. (p < 0.01), a total of 6537, according to the variance analysis results of the data set map volcano (Fig 3A).

**Fig. 3.**
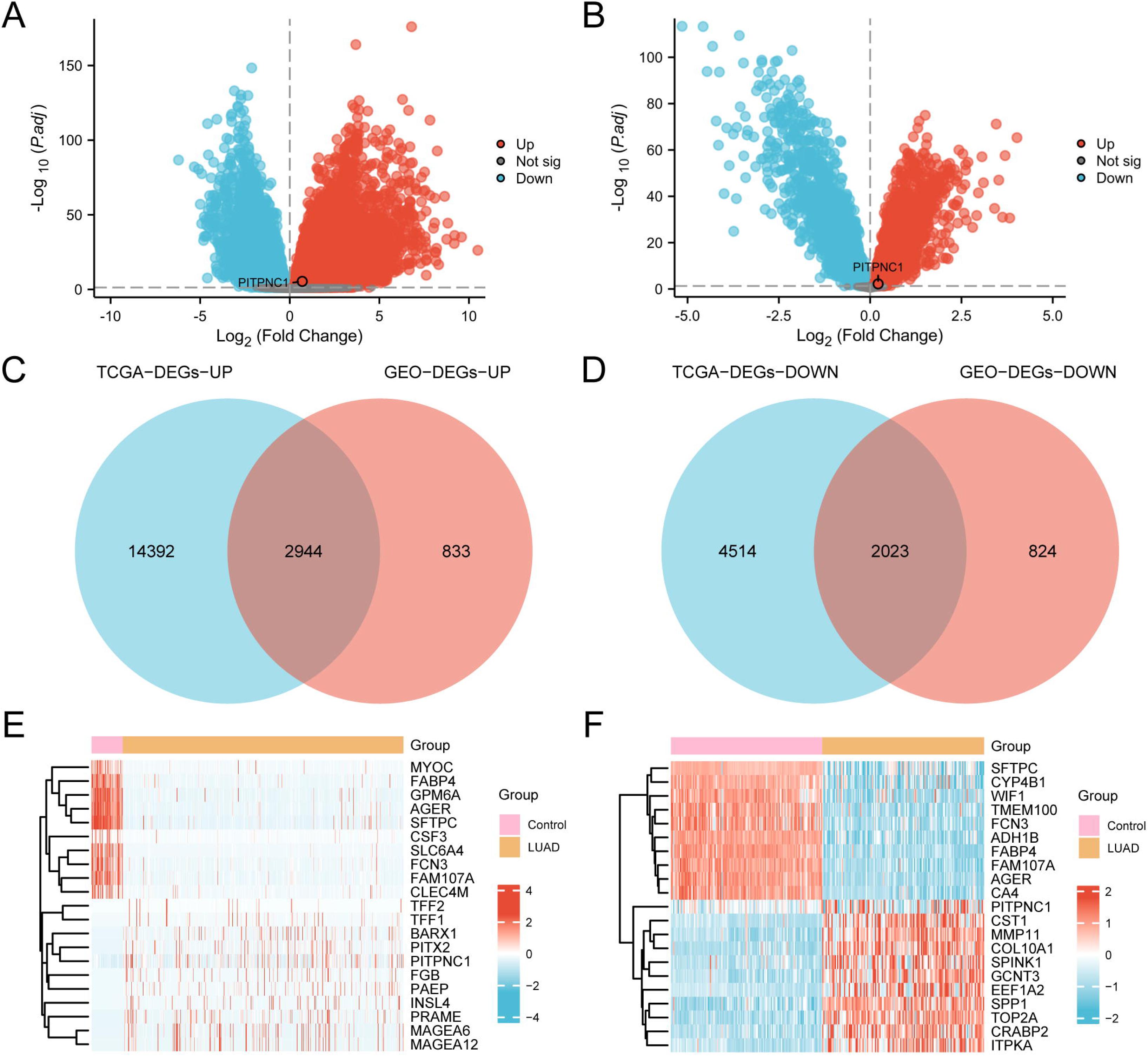
Differential Gene Expression Analysis. A. Volcano plot of differentially expressed genes analysis between lung adenocarcinoma (LUAD) group and Control (Control) group in the lung adenocarcinoma dataset (TCGA-LUAD). B. Volcano plot of differentially expressed gene analysis between lung adenocarcinoma (LUAD) group and Control (Control) group in the Combined Datasets. C. Venn diagram of up-regulated differentially expressed genes (DEGs) in the lung adenocarcinoma dataset (TCGA-LUAD) and the integrated GEO dataset (Combined Datasets). D. Down-regulated differentially expressed genes (DEGs) Venn diagram in lung adenocarcinoma Datasets (TCGA-LUAD) and integrated GEO datasets (Combined Datasets). E. logFC ranked TOP10 up-regulated and TOP10 down-regulated differentially expressed genes (DEGs) and heatmap of PITPNC1 in the lung adenocarcinoma dataset (TCGA-LUAD). F. Heat map of logFC-ranked TOP10 up-regulated and TOP10 down-regulated differentially expressed genes (DEGs) and PITPNC1 in Combined GEO Datasets. TCGA, The Cancer Genome Atlas; LUAD, Lung Adenocarcinoma; DEGs, Differentially Expressed Genes. In the heat map grouping, pink is the Control group, and orange is the lung adenocarcinoma (LUAD) group. In the heat map, red represents high expression and blue represents low expression.

Then, the data of the Combined GEO Datasets were divided into LUAD group and Control group. In order to examine the variations in gene expression levels between the LUAD group and the Control group within the Combined GEO Datasets, the R package limma was employed for the differential analysis. This approach facilitated the identification of differentially expressed genes across the two datasets, yielding the following results:

Combined Datasets a total of 6624 meet | logFC | > 0 and adj. P < 0.01 threshold of DEGs; Under the threshold, raised expressed genes (logFC > 0 and adj. P < 0.01), a total of 3777 cut expressed genes logFC < 0 and adj. (p < 0.01), a total of 2847, according to the variance analysis results of the data set map volcano (Fig 3B).

In order to obtain LUAD related DEGs, the intersection of all up-regulated DEGs with logFC > 0 and adj.p < 0.01 obtained from the TCGA-LUAD and the GEO dataset (Combined Datasets) was taken and the Venn diagram (Fig 3C) was drawn. Then, the intersection of all down-regulated DEGs with logFC < 0 and adj.p < 0.01 obtained from the TCGA-LUAD and the Combined Datasets was taken and the Venn diagram (Fig 3D) was drawn. A total of 4967 LUAD related DEGs were obtained after merging the intersection genes, and the detailed information is shown in TableS1. According to the intersection results, The TCGA-LUAD and the integrated GEO dataset (Combined The logFC sorted TOP10 up-regulated and TOP10 down-regulated DEGs and the expression difference of PITPNC1 in Datasets) were analyzed and the R package pheatmap was used to draw a heatmap to display the analysis results (Fig 3E-F).

### 3.4 Differential expression verification and ROC curve analysis of PITPNC1

To explore the expression differences of PITPNC1 in TCGA-LUAD and the Combined GEO Datasets, The difference analysis results of PITPNC1 expression in LUAD group and the Control group in the TCGA-LUAD (Fig 4A) and the Combined Datasets (Fig 4B) were shown by group comparison plots. The differential results showed that the expression of PITPNC1 in the LUAD group and the Control group of the TCGA-LUAD was highly statistically significant (p < 0.001). The expression of PITPNC1 in the LUAD group and the Control group of the Combined Datasets was highly statistically significant (p < 0.01). Finally, the R package pROC was used to draw ROC curves based on the expression of the gene PITPNC1 in the TCGA-LUAD (Fig 4C) and the Combined Datasets (Fig 4D). ROC curve showed that the expression level of PITPNC1 in the TCGA-LUAD had a low accuracy in the classification of LUAD group and Control group (0.5 < AUC < 0.7). The expression level of PITPNC1 in the Combined Datasets showed a low accuracy (0.5 < AUC < 0.7) in the classification of LUAD and Control groups.

**Fig. 4.**
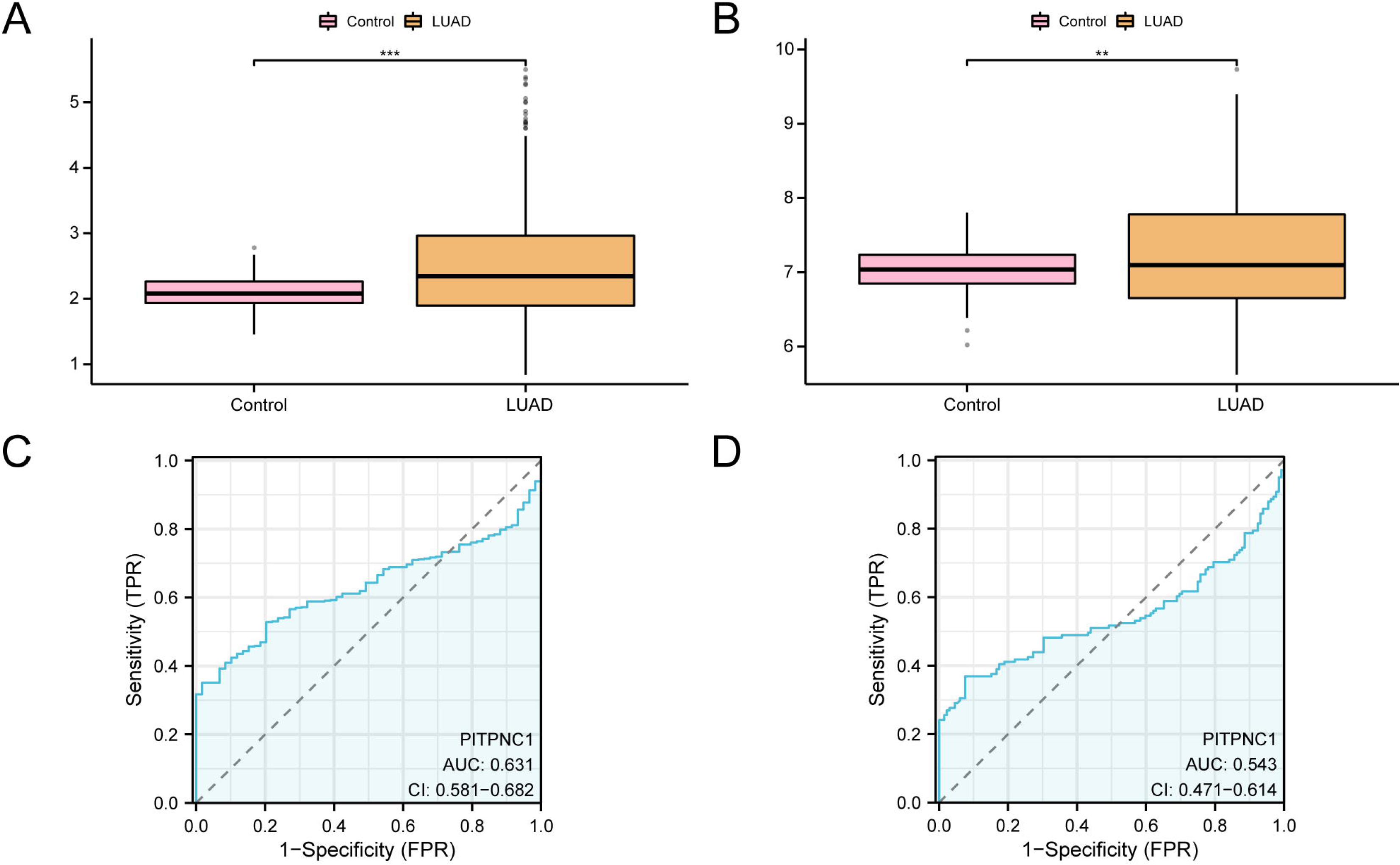
PPI Network and Differential Expression Validation and ROC Curve Analysis. A. Group comparison diagram of PITPNC1 gene in the lung adenocarcinoma (LUAD) group and the Control (Control) group of the lung adenocarcinoma dataset (TCGA-LUAD). B. Group comparison plot of PITPNC1 gene in the lung adenocarcinoma (LUAD) group and Control (Control) group of the Combined GEO Datasets. C. ROC curve of PITPNC1 in the lung adenocarcinoma dataset (TCGA-LUAD). D. ROC curves of PITPNC1 gene in the Combined GEO Datasets. TCGA, The Cancer Genome Atlas; LUAD, Lung Adenocarcinoma; ROC, Receiver Operating Characteristic; AUC, Area Under the Curve; TPR, True Positive Rate; FPR, False Positive Rate. In the group comparison figure, pink is the Control (Control) group and orange is the lung adenocarcinoma (LUAD) group. ** represents a p-value < 0.01, indicating a high degree of statistical significance; *** represents p value < 0.001 and highly statistically significant. When AUC > 0.5, it indicates that the expression of the molecule is a trend to promote the occurrence of the event, and the closer the AUC is to 1, the better the diagnostic effect. AUC values between 0.5 and 0.7 were associated with lower accuracy.

### 3.5 Differential analysis and co-expressed genes of PITPNC1 high and low expression groups

Firstly, the LUAD samples of the TCGA-LUAD were divided into high expression group and low expression group according to the median expression of PITPNC1. To analyze the difference of gene expression values between high expression group and low expression group in the LUAD samples of the TCGA-LUAD, The R package DESeq2 was used for differential analysis of LUAD samples in the TCGA-LUAD to obtain the differentially expressed genes in the two groups of data, and the results are as follows: TCGA-LUAD datasets specimens of 3838 patients with LUAD meet |logFC | > 1.5 and adj. P < 0.01 threshold of DEGs; Under the threshold, raised expressed genes (logFC > 1.5 and adj., p < 0.01), a total of 273 cut expressed genes (logFC < - 1.5 and adj. P < 0.01), a total of 3565, according to the results of variance analysis map volcano (Fig 5A). Then, the logFC ranked TOP10 up-regulated and TOP10 down-regulated DEGs and PITPNC1 expression differences in the LUAD samples of the TCGA-LUAD were analyzed, and the R package pheatmap was used to draw a heatmap to display the analysis results (Fig 5B).

**Fig. 5.**
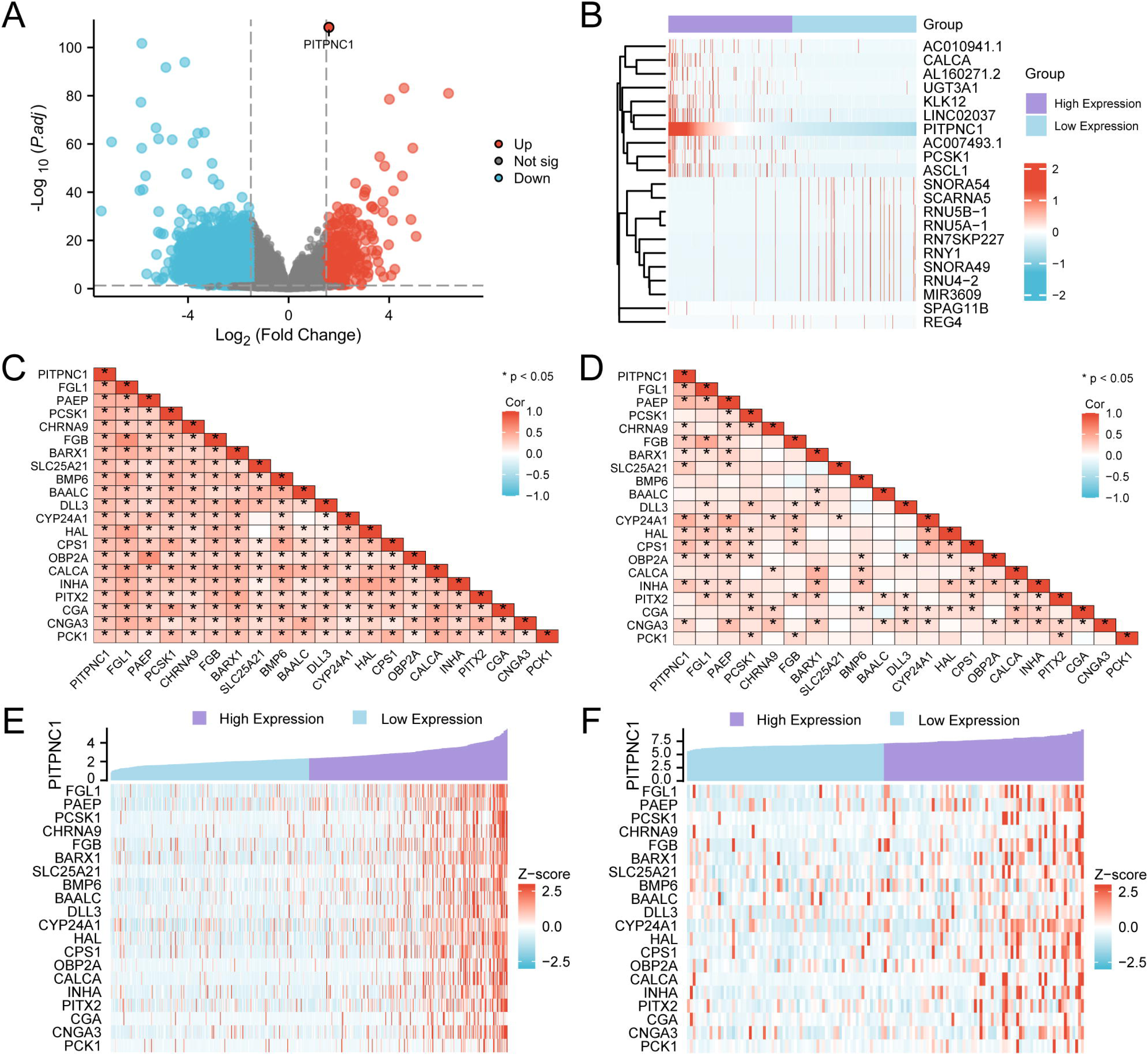
Expression Difference Analysis of Expression Group of PITPNC1. A. Volcano plot of differential expression between PITPNC1 high and low expression groups in lung adenocarcinoma (LUAD) samples from the Lung Adenocarcinoma dataset (TCGA-LUAD). B. logFC ranked TOP10 up-regulated and TOP10 down-regulated differentially expressed genes (DEGs) and heat map of PITPNC1 in lung adenocarcinoma (LUAD) samples from the Lung Adenocarcinoma dataset (TCGA-LUAD). C-d. Correlation heatmap between PITPNC1 and Co-expressed Genes in lung adenocarcinoma (LUAD) samples from TCGA-LUAD Datasets (C) and Combined GEO datasets (D). E-f. PITPNC1 and Co-expressed genes in PITPNC1 high and low expression groups in lung adenocarcinoma (LUAD) samples from the TCGA-LUAD dataset (E) and the integrated GEO Datasets (F) Genes) co-expression heatmap. TCGA, The Cancer Genome Atlas; LUAD, Lung Adenocarcinoma; DEGs, Differentially Expressed Genes. The absolute value of correlation coefficient (r value) below 0.3 was weak or no correlation, 0.3-0.5 was weak correlation, and 0.5-0.8 was moderate correlation. * represents p value < 0.05, indicating statistical significance. In the correlation heat map, red indicates positive correlation and blue indicates negative correlation. Red is positive correlation and blue is negative correlation in the correlation heat map. In the co-expression heatmap group, purple is the High Expression group, and blue is the Low Expression group. In the co-expression heatmap, red is high expression and blue is low expression.

Then, the DEGs from the high-low expression group differential analysis and all genes in the LUAD samples of the Combined Datasets were intersected to obtain 133 LUAD related DEGs. For detailed information, see TableS2. Then the correlation analysis was performed between PITPNC1 and other LUAD related DEGs. The top 20 genes with the largest absolute value of correlation coefficient (r value) were Co-expressed genes, which were FGL1, PAEP, PCSK1, and LUAD. CHRNA9, FGB, BARX1, SLC25A21, BMP6, BAALC, DLL3, CYP24A1, HAL, CPS1, OBP2A, CALCA, INHA, PITX2, CGA, CNGA3, PCK1, and the correlation heat map was drawn (Fig 5C). The results showed that PITPNC1 gene was positively correlated with all Co-expressed Genes (r value > 0). Then, the correlation between PITPNC1 gene and 20 Co-expressed Genes in the LUAD samples of the Combined GEO Datasets was analyzed, and the correlation heat map was drawn (Fig 5D). The results showed that PITPNC1 was positively correlated with all Co-expressed Genes (r value > 0). Finally, the co-expression heatmap of 20 Co-expressed Genes and PITPNC1 was drawn (Fig 5E-F).

### 3.6 Gene Ontology (GO) and pathway (KEGG) enrichment analysis

GO and KEGG enrichment analysis were used to further explore the relationship between biological process (BP), cellular component (CC), molecular function (MF) and biological pathway (KEGG) of 133 DEGs and LUAD. These 133 DEGs were used for GO and KEGG enrichment analysis, and the specific results are shown in Table 3. The results showed that 133 DEGs were mainly enriched in response to xenobiotic stimulus, regulation of hormone levels, and response to xenobiotic stimulus in LUAD. endocrine system development, cellular response to glucagon stimulus and neuropeptide signaling pathway and other biological processes (BP); neuronal cell body, axon terminus, neuron projection terminus, terminal bouton and distal axon (CC); hormone activity, receptor ligand activity, signaling receptor activator activity, carbonate dehydratase activity and metal ion transmembrane transporter activity in molecular function (MF). At the same time, it was also enriched in Nitrogen metabolism and Neuroactive ligand-receptor interaction biological pathway. The results of GO and pathway enrichment analysis were visualized by bar diagram (Fig 6A) and bubble diagram (Fig 6B).

**Fig. 6.**
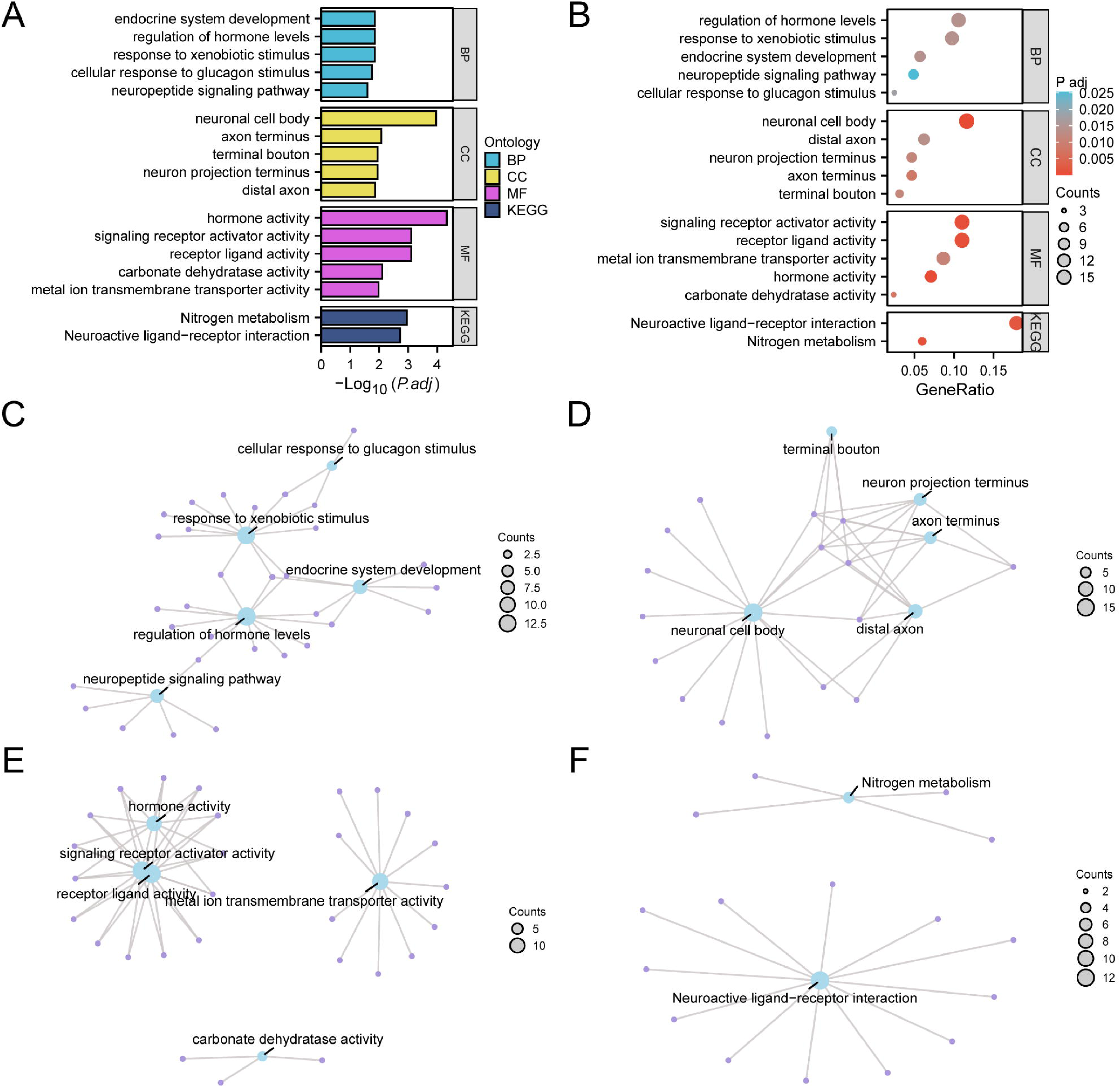
GO and KEGG Enrichment Analysis for DEGs. A-b. Gene ontology (GO) and pathway (KEGG) enrichment analysis results of differentially expressed genes (DEGs) Bar graph (A) and bubble plot (B) show: biological process (BP), cellular component (CC), molecular function (MF) and biological pathway (KEGG). GO terms and KEGG terms are shown on the ordinate. C-f. Gene ontology (GO) and pathway (KEGG) enrichment analysis results network diagram of differentially expressed genes (DEGs) : BP (C), CC (D), MF (E) and KEGG (F). Blue nodes represent items, purple nodes represent molecules, and lines represent the relationship between items and molecules. DEGs, Differentially Expressed Genes; GO, Gene Ontology; KEGG, Kyoto Encyclopedia of Genes and Genomes; BP, Biological Process; CC, Cell Component; MF, Molecular Function. The bubble size in the bubble plot represents the number of genes, and the color of the bubble represents the size of the adj. P-value, the reder the color, the smaller the adj. P-value, and the bluer the color, the larger the adj. P-value. The screening criteria for gene ontology (GO) and pathway (KEGG) enrichment analysis were adj.p < 0.05 and FDR value (q value) < 0.25, and the p value correction method was Benjamini-Hochberg (BH).

**Table 3.**
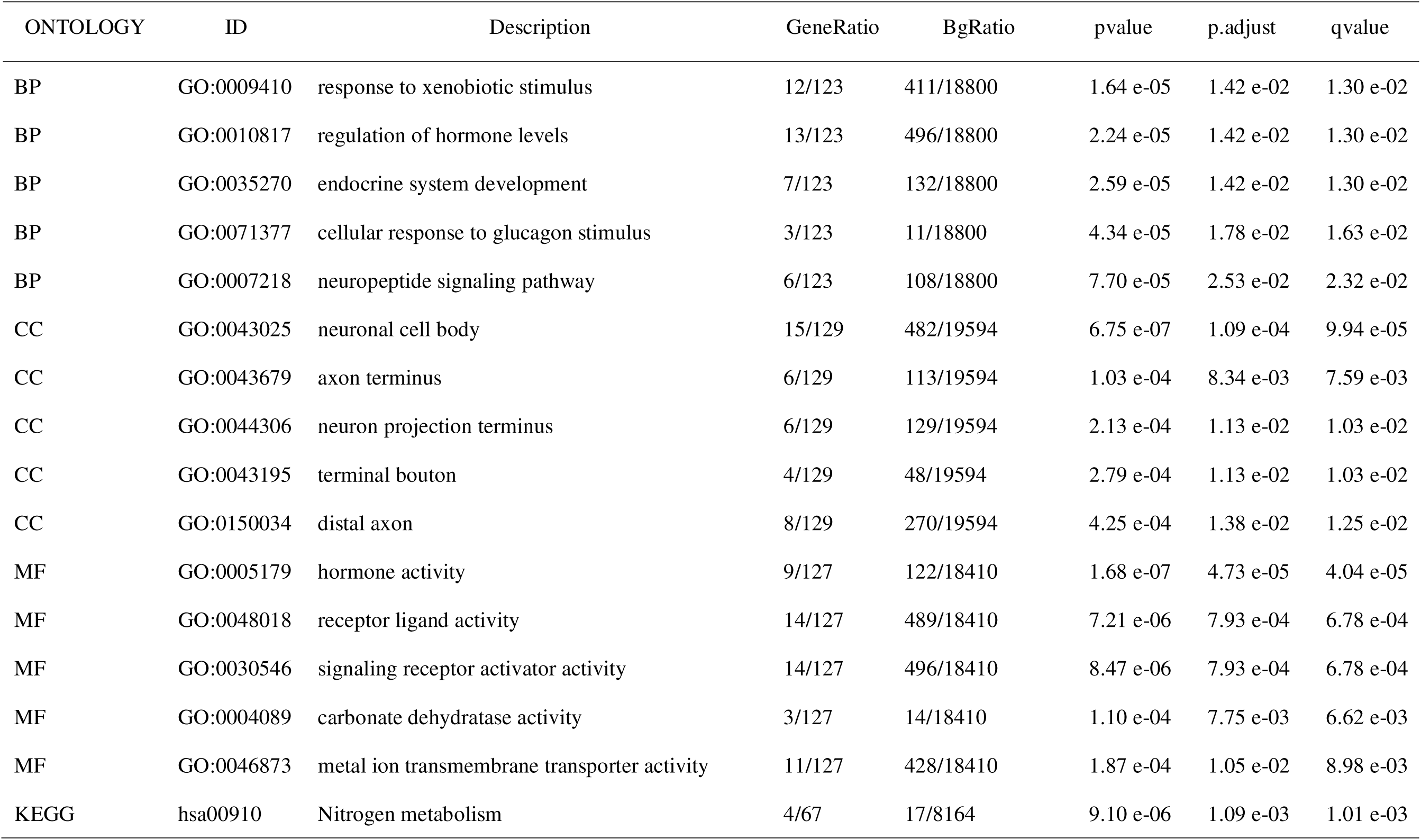
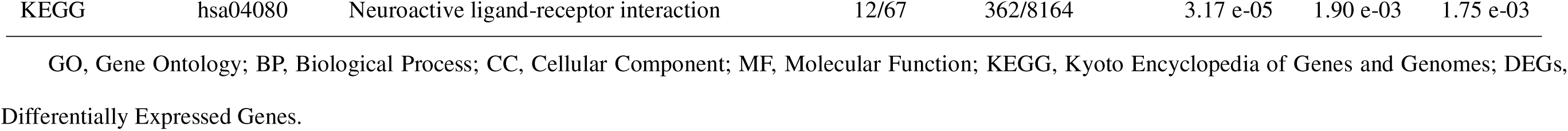
Result of GO and KEGG Enrichment Analysis for DEGs.

| ONTOLOGY | ID | Description | GeneRatio | BgRatio | pvalue | p.adjust | qvalue |
| --- | --- | --- | --- | --- | --- | --- | --- |
| BP | GO:0009410 | response to xenobiotic stimulus | 12/123 | 411/18800 | 1.64 e-05 | 1.42 e-02 | 1.30 e-02 |
| BP | GO:0010817 | regulation of hormone levels | 13/123 | 496/18800 | 2.24 e-05 | 1.42 e-02 | 1.30 e-02 |
| BP | GO:0035270 | endocrine system development | 7/123 | 132/18800 | 2.59 e-05 | 1.42 e-02 | 1.30 e-02 |
| BP | GO:0071377 | cellular response to glucagon stimulus | 3/123 | 11/18800 | 4.34 e-05 | 1.78 e-02 | 1.63 e-02 |
| BP | GO:0007218 | neuropeptide signaling pathway | 6/123 | 108/18800 | 7.70 e-05 | 2.53 e-02 | 2.32 e-02 |
| CC | GO:0043025 | neuronal cell body | 15/129 | 482/19594 | 6.75 e-07 | 1.09 e-04 | 9.94 e-05 |
| CC | GO:0043679 | axon terminus | 6/129 | 113/19594 | 1.03 e-04 | 8.34 e-03 | 7.59 e-03 |
| CC | GO:0044306 | neuron projection terminus | 6/129 | 129/19594 | 2.13 e-04 | 1.13 e-02 | 1.03 e-02 |
| CC | GO:0043195 | terminal bouton | 4/129 | 48/19594 | 2.79 e-04 | 1.13 e-02 | 1.03 e-02 |
| CC | GO:0150034 | distal axon | 8/129 | 270/19594 | 4.25 e-04 | 1.38 e-02 | 1.25 e-02 |
| MF | GO:0005179 | hormone activity | 9/127 | 122/18410 | 1.68 e-07 | 4.73 e-05 | 4.04 e-05 |
| MF | GO:0048018 | receptor ligand activity | 14/127 | 489/18410 | 7.21 e-06 | 7.93 e-04 | 6.78 e-04 |
| MF | GO:0030546 | signaling receptor activator activity | 14/127 | 496/18410 | 8.47 e-06 | 7.93 e-04 | 6.78 e-04 |
| MF | GO:0004089 | carbonate dehydratase activity | 3/127 | 14/18410 | 1.10 e-04 | 7.75 e-03 | 6.62 e-03 |
| MF | GO:0046873 | metal ion transmembrane transporter activity | 11/127 | 428/18410 | 1.87 e-04 | 1.05 e-02 | 8.98 e-03 |
| KEGG | hsa00910 | Nitrogen metabolism | 4/67 | 17/8164 | 9.10 e-06 | 1.09 e-03 | 1.01 e-03 |
| KEGG | hsa04080 | Neuroactive ligand-receptor interaction | 12/67 | 362/8164 | 3.17 e-05 | 1.90 e-03 | 1.75 e-03 |

GO, Gene Ontology; BP, Biological Process; CC, Cellular Component; MF, Molecular Function; KEGG, Kyoto Encyclopedia of Genes and Genomes; DEGs, Differentially Expressed Genes.

Concurrently, a network diagram illustrating the biological process (BP), cellular component (CC), molecular function (MF), and biological pathway (KEGG) was constructed based on the findings from GO and KEGG enrichment analyses (Fig. 6C-F). The connecting lines represent the associated molecules along with their respective annotations, while the size of the nodes indicates the quantity of molecules encompassed within each entry. The analysis revealed a notable enrichment of genes within the neuronal cell body pertaining to the cellular component (CC).

### 3.7 Gene set Enrichment analysis (GSEA)

To determine the influence of the expression levels of all genes in LUAD samples on the risk of developing LUAD, GSEA was used to investigate the relationship between the expression levels of all genes in LUAD samples and the biological processes, cellular components and molecular functions they play. And presented by bubble plot (Fig 7A), the specific results are shown in Table 4. The results showed that all genes in LUAD samples were significantly enriched in IL12 2pathway (Fig 7B), Signaling By Notch (Fig 7C), MAPK6, MAPK4 Signaling (Fig 7D), Hedgehog On State (Fig 7E) and other biologically relevant functions and signaling pathways.

**Fig. 7.**
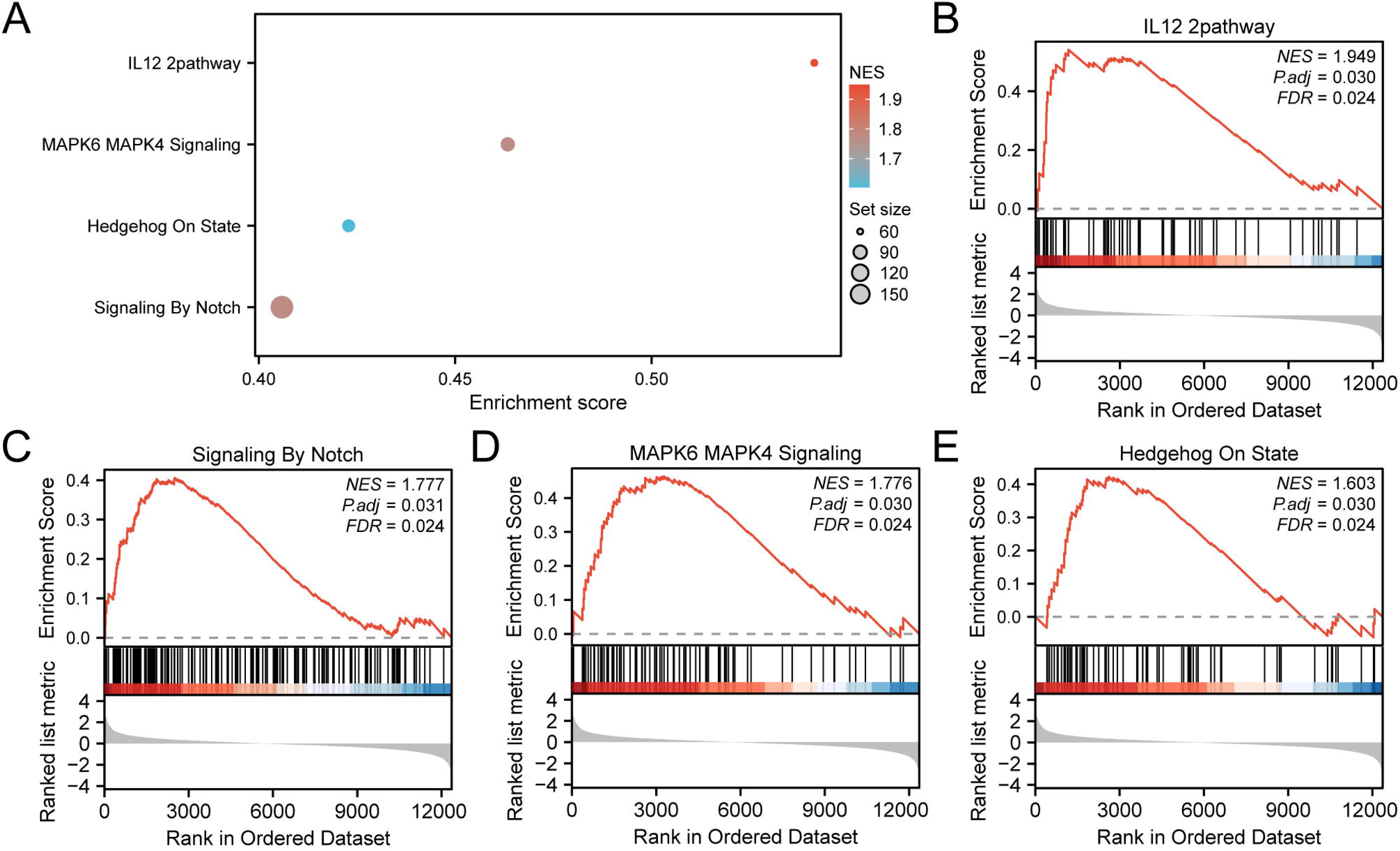
GSEA for Risk Group. A. Bubble plot presentation of 4 biological functions of gene set enrichment analysis (GSEA) of lung adenocarcinoma (LUAD) samples. Gene set enrichment analysis (GSEA) showed that all genes were significantly enriched in IL12 2pathway (B), Signaling By Notch (C), MAPK6 MAPK4 Signaling (D) and Hedgehog On State (E). LUAD, Lung Adenocarcinoma; GSEA, Gene Set Enrichment Analysis. In the bubble plot, the size of the bubble represents the number of enriched genes, and the color of the bubble represents the size of the NES value. The more red the color, the larger the NES, and the more blue the smaller the NES. The screening criteria of gene set enrichment analysis (GSEA) were adj.p < 0.05 and FDR value (q value) < 0.25, and the p value correction method was Benjamini-Hochberg (BH).

**Table 4.** Results of GSEA for Expression Group.

| ID | Set Size | Enrichment<br>Score | NES | p value | p.adjust | q value |
| --- | --- | --- | --- | --- | --- | --- |
| REACTOME_CELL_CYCLE_CHECKPOINTS | 237 | 6.67 e-01 | 3.04 e+00 | 3.31 e-03 | 3.19 e-02 | 2.54 e-02 |
| REACTOME_MITOTIC_G1_PHASE_AND_G1_S_TRANSITION | 142 | 7.01 e-01 | 3.00 e+00 | 2.84 e-03 | 3.05 e-02 | 2.43 e-02 |
| REACTOME_DNA_REPLICATION | 137 | 6.96 e-01 | 2.96 e+00 | 2.78 e-03 | 3.04 e-02 | 2.42 e-02 |
| REACTOME_CELL_CYCLE_MITOTIC | 458 | 6.07 e-01 | 2.95 e+00 | 4.65 e-03 | 3.91 e-02 | 3.12 e-02 |
| REACTOME_G2_M_CHECKPOINTS | 134 | 6.87 e-01 | 2.90 e+00 | 2.76 e-03 | 3.04 e-02 | 2.42 e-02 |
| REACTOME_SYNTHESIS_OF_DNA | 110 | 7.11 e-01 | 2.90 e+00 | 2.72 e-03 | 3.04 e-02 | 2.42 e-02 |
| REACTOME_MITOTIC_METAPHASE_AND_ANAPHASE | 201 | 6.50 e-01 | 2.89 e+00 | 3.21 e-03 | 3.16 e-02 | 2.52 e-02 |
| WP_RETINOBLASTOMA_GENE_IN_CANCER | 84 | 7.30 e-01 | 2.81 e+00 | 2.62 e-03 | 3.04 e-02 | 2.42 e-02 |
| REACTOME_S_PHASE | 145 | 6.55 e-01 | 2.80 e+00 | 2.92 e-03 | 3.06 e-02 | 2.44 e-02 |
| REACTOME_MITOTIC_SPINDLE_CHECKPOINT | 92 | 7.02 e-01 | 2.77 e+00 | 2.61 e-03 | 3.04 e-02 | 2.42 e-02 |
| REACTOME_DNA_REPLICATION_PRE_INITIATION | 111 | 6.81 e-01 | 2.76 e+00 | 2.82 e-03 | 3.04 e-02 | 2.42 e-02 |
| REACTOME_SEPARATION_OF_SISTER_CHROMATIDS | 162 | 6.43 e-01 | 2.76 e+00 | 3.06 e-03 | 3.07 e-02 | 2.45 e-02 |
| KEGG_CELL_CYCLE | 114 | 6.69 e-01 | 2.73 e+00 | 2.81 e-03 | 3.04 e-02 | 2.42 e-02 |
| REACTOME_ACTIVATION_OF_THE_PRE_REPLICATIVE_COMPLEX | 31 | 8.42 e-01 | 2.69 e+00 | 2.40 e-03 | 3.04 e-02 | 2.42 e-02 |
| WP_CELL_CYCLE | 111 | 6.63 e-01 | 2.69 e+00 | 2.82 e-03 | 3.04 e-02 | 2.42 e-02 |
| REACTOME_RESOLUTION_OF_SISTER_CHROMATID_COHESION | 106 | 6.66 e-01 | 2.68 e+00 | 2.76 e-03 | 3.04 e-02 | 2.42 e-02 |
| PID_IL12_2PATHWAY | 60 | 5.41 e-01 | 1.95 e+00 | 2.54 e-03 | 3.04 e-02 | 2.42 e-02 |
| REACTOME_SIGNALING_BY_NOTCH | 178 | 4.06 e-01 | 1.78 e+00 | 3.02 e-03 | 3.06 e-02 | 2.44 e-02 |
| REACTOME_MAPK6_MAPK4_SIGNALING | 82 | 4.63 e-01 | 1.78 e+00 | 2.60 e-03 | 3.04 e-02 | 2.42 e-02 |
| REACTOME_HEDGEHOG_ON_STATE | 74 | 4.23 e-01 | 1.60 e+00 | 2.53 e-03 | 3.04 e-02 | 2.42 e-02 |

GSEA, Gene Set Enrichment Analysis.

### 3.8 Construction of protein-protein interaction Network (PPI Network)

The interaction network of PITPNC1 and its functional similar genes was predicted by GeneMANIA website (Fig 8), and the lines with different colors represent the co-expression between them, sharing protein domains and other information. Among them, it contains gene PITPNC1 and 20 functionally similar proteins, and the detailed information is shown in TableS3.

**Fig. 8.**
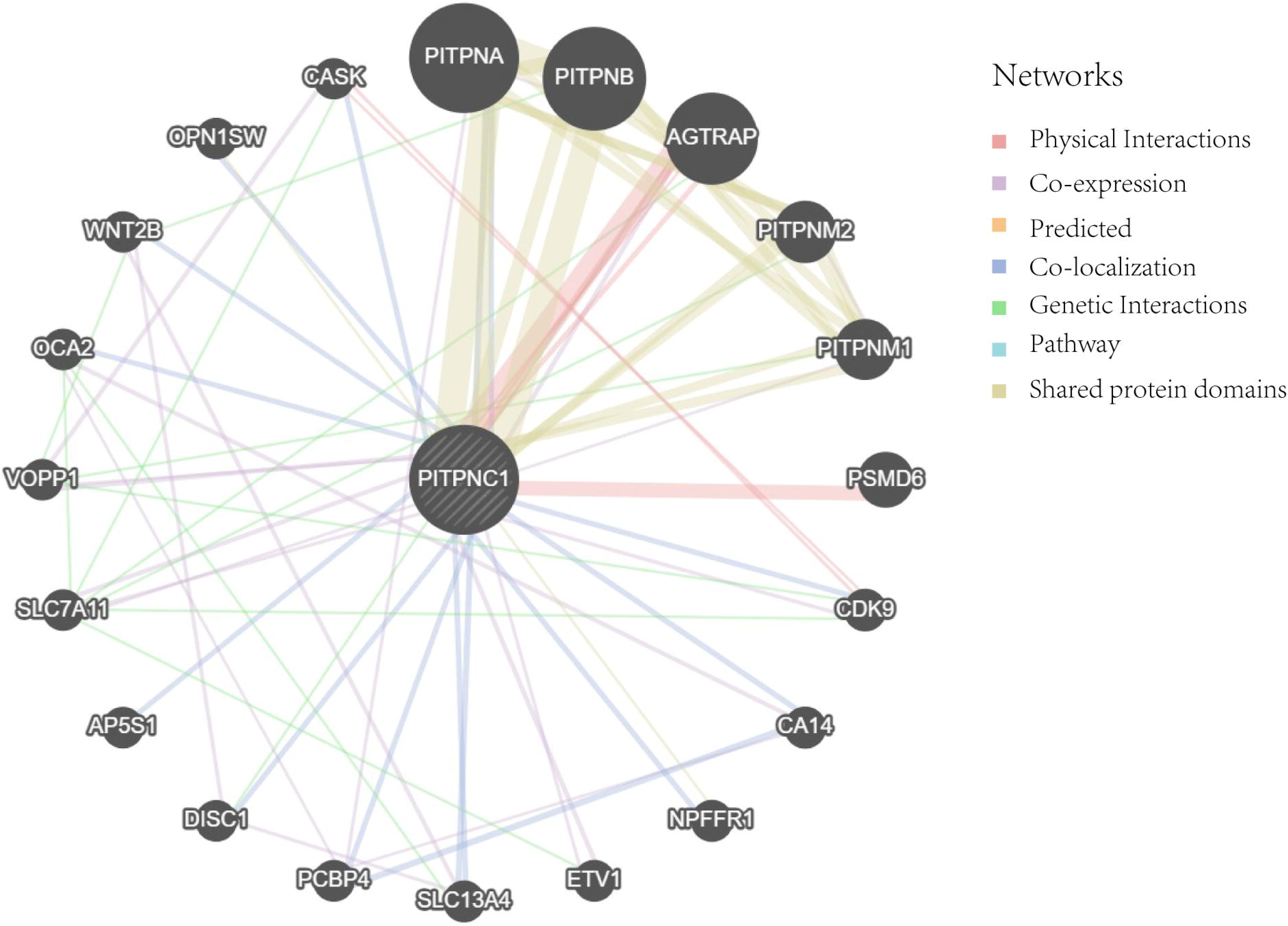
PPI Network Analysis. The protein-protein interaction Network (PPI Network) of PITPNC1 was predicted by GeneMANIA website. The circles in the figure show PITPNC1 and its functionally similar genes, and the colors corresponding to the lines represent the interconnected functions. PPI, Protein-protein Interaction.

### 3.9 Construction of regulatory network

Firstly, the transcription factors (TFS) that bind to PITPNC1 were obtained from ChIPBase database, and the mRNA-TF Regulatory Network was constructed and visualized using Cytoscape software (Fig 9A). Among them, it contains gene PITPNC1 and 17 transcription factors, and the detailed information is shown in TableS4.

**Fig. 9.**
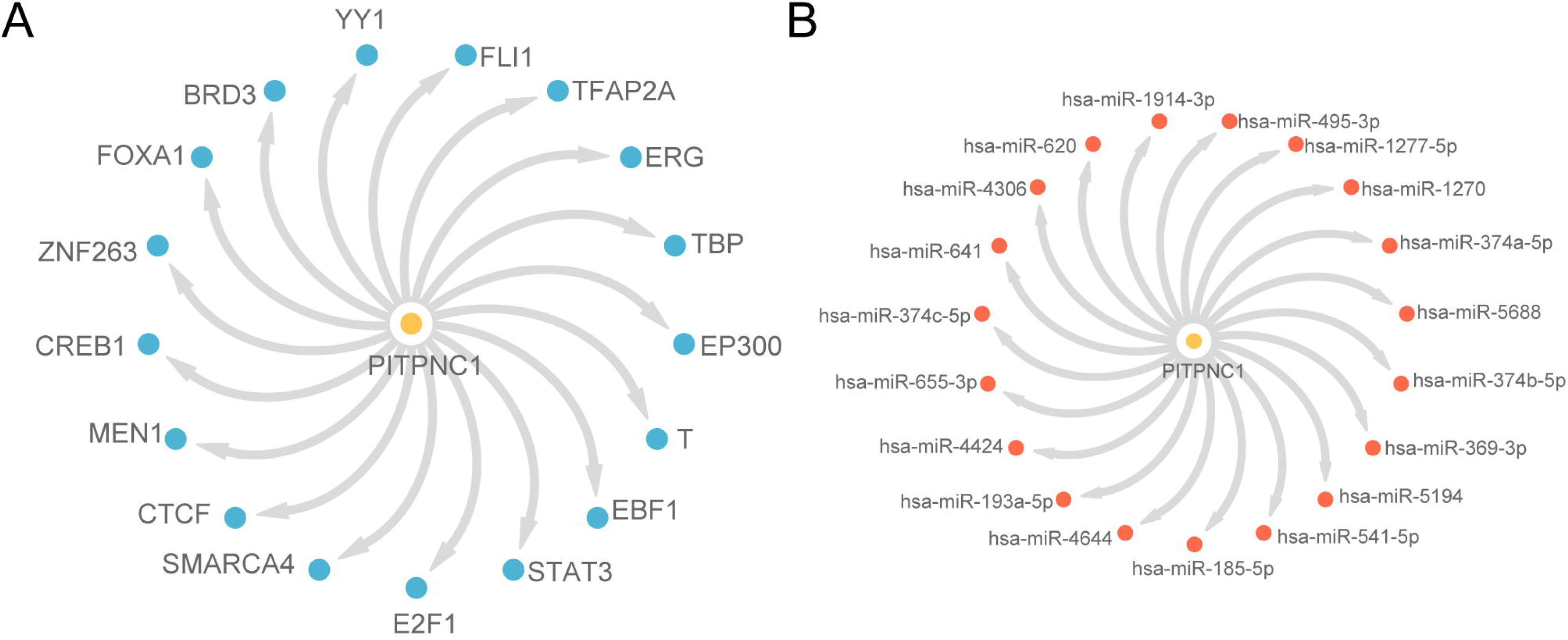
Regulatory Network of PITPNC1. A. The mRNA-TF Regulatory Network of PITPNC1. B. mRNA-miRNA Regulatory Network of PITPNC1. TF, Transcription Factor. Yellow is mRNA, blue is TF, and red is miRNA.

Then, the miRNA related to PITPNC1 was obtained from StarBase database, and the mRNA-miRNA Regulatory Network was constructed and visualized by Cytoscape software (Fig 9B). Among them, the gene PITPNC1 and 19 miRNAs were included, and the detailed information is shown in TableS5.

### 3.10 Immune infiltration analysis of high and low expression groups (ssGSEA)

The expression matrix of LUAD samples in the TCGA-LUAD was utilized to calculate the immune infiltration abundance of 28 immune cells in LUAD samples by ssGSEA algorithm. Firstly, the expression differences of infiltrating abundance of immune cells in different groups were shown by group comparison plot. The group comparison diagram (Fig10A) showed that all the five immune cells were statistically significant (p < 0.05), which were: CD56dim natural killer cell, Eosinophil, Neutrophil, Type 17 T helper cell, Type 2 T helper cell. Then, the correlation results of the abundance of five immune cell infiltration in LUAD samples were shown by correlation heat map (Fig 10B-C). The results showed that most of the immune cells in the high expression group of LUAD samples showed strong positive correlation, and the immune cell Type 2 T helper cell and Neutrophil had the strongest significant positive correlation (r = 0.459, P < 0.05). Most of the immune cells in the Low Expression group showed strong positive correlation, and the immune cell Type 17 T helper cell and Neutrophil had the strongest significant positive correlation (r = 0.464, p < 0.05). Finally, the correlation between PITPNC1 gene and the abundance of immune cell infiltration was shown by correlation bubble plot (Fig 10D-E). The results of the correlation bubble plot showed that: Most of the immune cells in the high expression group of LUAD samples showed strong positive correlation, and the gene PITPNC1 and immune cell CD56dim natural killer cell had the strongest significant positive correlation (r = 0.237, P < 0.05). Most of the immune cells in the low expression group showed strong correlation, and the gene PITPNC1 and immune cell Type 2 T helper cell had the strongest significant positive correlation (r = 0.19, p < 0.05).

**Fig. 10.**
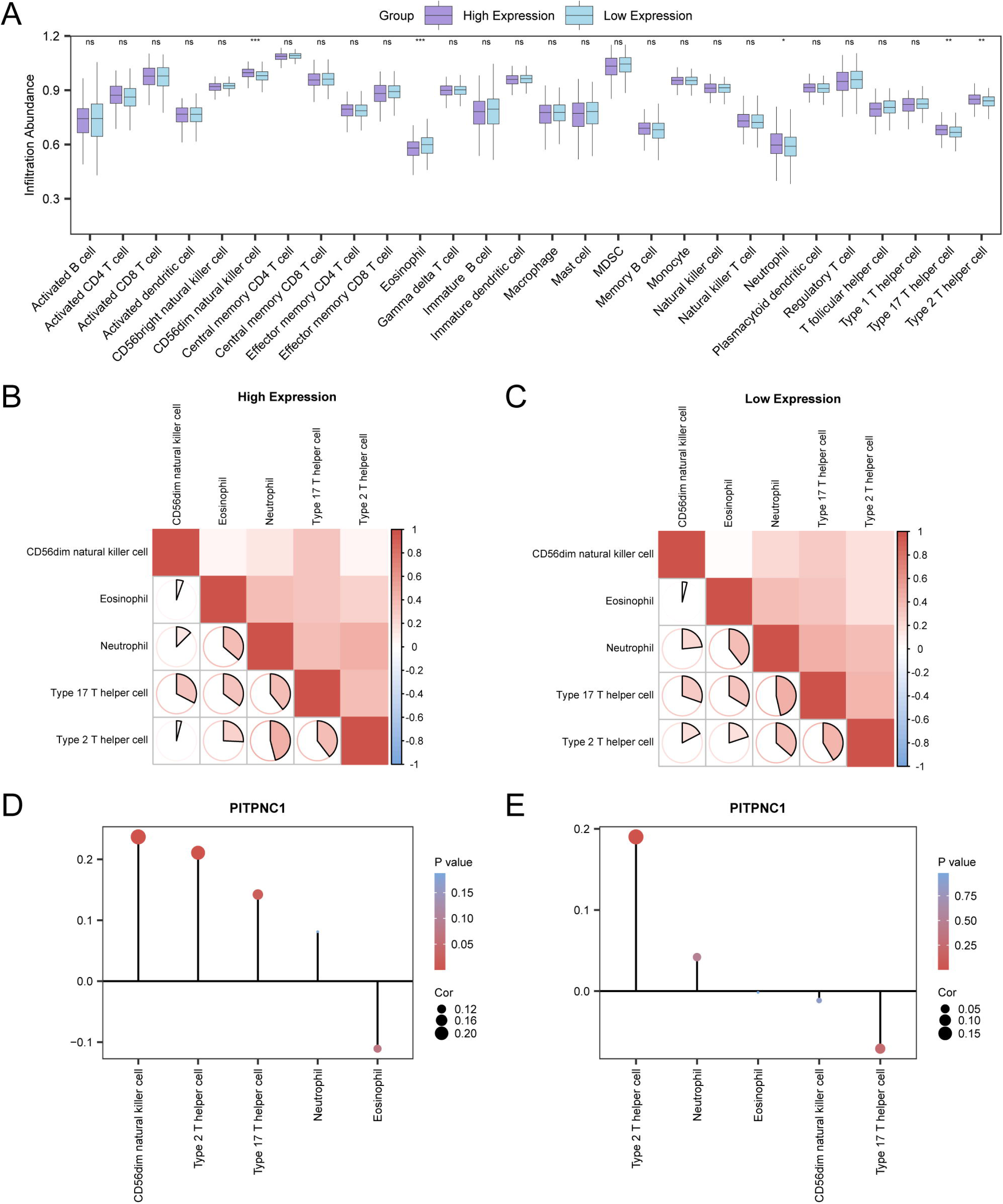
Risk Group Immune Infiltration Analysis by ssGSEA Algorithm. A. Comparison of the grouping of immune cells in the High Expression group and Low Expression group of lung adenocarcinoma (LUAD) samples. B-c. Results of correlation analysis of immune cell infiltration abundance in High Expression group (B) and Low Expression group (C) of lung adenocarcinoma (LUAD) samples are shown. D-e. Bubble plot of correlation between immune cell infiltration abundance and PITPNC1 in High Expression group (D) and Low Expression group (E) of lung adenocarcinoma (LUAD) samples. ssGSEA, single-sample Gene-Set Enrichment Analysis; LUAD, Lung Adenocarcinoma. ns stands for p value ≥ 0.05, not statistically significant; * represents p value < 0.05, statistically significant; ** represents p value < 0.01, highly statistically significant; *** represents p value < 0.001 and highly statistically significant. The absolute value of the correlation coefficient (r value) below 0.3 was considered as weak or no correlation, and the r value between 0.3 and 0.5 was considered as weak correlation. Purple is the High Expression group, blue is the Low Expression group. Red is positive correlation, blue is negative correlation, and the depth of the color represents the strength of the correlation.

### 3.11 Construction of clinical prognostic model and prognostic analysis of lung adenocarcinoma

Firstly, time-dependent ROC curves (Fig 11A) were plotted for the LUAD samples from the TCGA-LUAD. The results showed that the clinical prognostic model of LUAD had low accuracy (0.7 > AUC > 0.5) at 1 year, 2 years and 3 years. In addition, we also performed prognostic Kaplan-Meier (KM) curve analysis based on PITPNC1 expression in LUAD samples combined with median grouping of overall survival (OS) in LUAD samples from the TCGA-LUAD dataset (Fig 11B). The results showed that there was a statistically significant difference in overall survival (OS) between the high expression group and the low expression group and the LUAD samples in the TCGA-LUAD (p < 0.05).

**Fig. 11.**
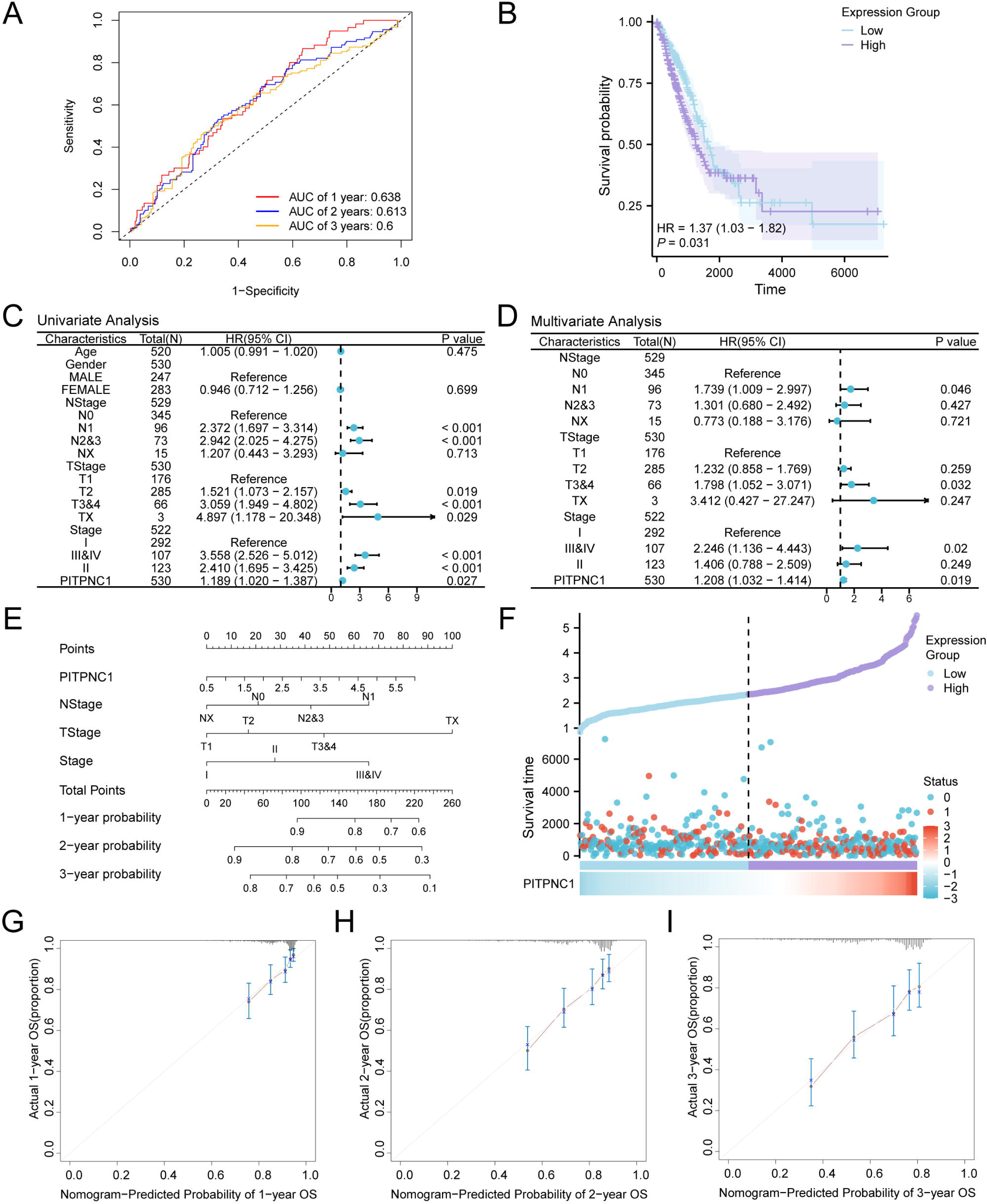
Prognostic Analysis. A. Time-dependent ROC curves of lung adenocarcinoma (LUAD) samples in the Lung adenocarcinoma dataset (TCGA-LUAD). B. Prognostic KM curve between PITPNC1 expression level and overall survival (OS) of LUAD samples. C. Forest Plot of PITPNC1 expression level and clinical information in univariate Cox regression model (C) and multivariate Cox regression model (D). E. Nomogram of PITPNC1 expression level and clinical information in univariate and multivariate Cox regression model. F. Risk factor map of lung adenocarcinoma (LUAD) samples in the Lung Adenocarcinoma Dataset (TCGA-LUAD). G-i. 1 year (G), 2 year (H), and 3 year (I) Calibration Curve of the prognostic risk model for lung adenocarcinoma (LUAD). TCGA, The Cancer Genome Atlas; LUAD, Lung Adenocarcinoma; OS, Overall Survival; KM, Kaplan-Meier; ROC, Receiver Operating Characteristic Curve; AUC, Area Under the Curve. When AUC > 0.5, it indicates that the expression of the molecule is a trend to promote the occurrence of the event, and the closer the AUC is to 1, the better the diagnostic effect. AUC values between 0.5-0.7 were associated with lower accuracy. p value < 0.05 was considered statistically significant.

Then, univariate Cox regression analysis was performed based on the median PITPNC1 expression level in LUAD samples combined with the overall survival (OS) and clinical information of LUAD samples, and variables with p < 0.10 were screened for multivariate Cox regression analysis. The results of univariate and multivariate Cox regression analysis were visualized by Forest Plot (Fig 11C-D), and comprehensive details are presented in Table 5. The outcomes derived from the univariate multivariate Cox regression analysis indicated that the expression level of PITPNC1 gene and clinical information NStage, TStage, Stage were statistically significant (p < 0.05).

**Table 5.** Results of Cox Analysis.

| Characteristics | Total(N) | Univariate analysis |  | Multivariate analysis |  |
| --- | --- | --- | --- | --- | --- |
|  |  | HR (95% CI) | P value | HR(95% CI) | P value |
| Age | 520 | 1.005 (0.991-1.020) | 0.475 |  |  |
| Gender | 530 |  |  |  |  |
| MALE | 247 | Reference |  |  |  |
| female | 283 | 0.946 (0.712-1.256) | 0.699 |  |  |
| NStage | 529 |  |  |  |  |
| N0 | 345 | Reference |  | Reference |  |
| N1 | 96 | 2.372 (1.697-3.314) | < 0.001 | 1.739 (1.009-2.997) | 0.046 |
| N2&3 | 73 | 2.942 (2.025-4.275) | < 0.001 | 1.301 (0.680-2.492) | 0.427 |
| NX | 15 | 1.207 (0.443-3.293) | 0.713 | 0.773 (0.188-3.176) | 0.721 |
| TStage | 530 |  |  |  |  |
| T1 | 176 | Reference |  | Reference |  |
| T2 | 285 | 1.521 (1.073-2.157) | 0.019 | 1.232 (0.858-1.769) | 0.259 |
| T3&4 | 66 | 3.059 (1.949-4.802) | < 0.001 | 1.798 (1.052-3.071) | 0.032 |
| TX | 3 | 4.897 (1.178-20.348) | 0.029 | 3.412 (0.427-27.247) | 0.247 |
| Stage | 522 |  |  |  |  |
| I | 292 | Reference |  | Reference |  |
| III&IV | 107 | 3.558 (2.526-5.012) | < 0.001 | 2.246 (1.136-4.443) | 0.020 |
| II | 123 | 2.410 (1.695-3.425) | < 0.001 | 1.406 (0.788-2.509) | 0.249 |
| PITPNC1 | 530 | 1.189 (1.020-1.387) | 0.027 | 1.208 (1.032-1.414) | 0.019 |

B. HR, Hazard ratio, general HR > 1 indicates that the variable is a risk factor, and HR < 1 is a protective factor.
Univariate p values < 0.1 were included in the analysis.

In order to further explore the value of clinical prognostic model of LUAD, a Nomogram was drawn based on the findings derived from both univariate and multivariate Cox regression analyses, it is essential to demonstrate the relationship between PITPNC1 expression level and three clinical information in LUAD samples (Fig 11E). The results showed that the utility of TStage in the clinical prognostic model of LUAD was significantly higher than that of other variables.

Subsequently, The R package ggplot2 was utilized to visualize the risk factors associated with the expression levels of the gene PITPNC1 (Fig 11F). The findings indicated that the number of mortality occurrences in the high expression cohort exceeded that of the low expression cohort.

Furthermore, we performed 1 -, 2 -, and 3-year prognostic Calibration analysis on the clinical prognostic model of LUAD and plotted a Calibration Curve (Fig 11G-I). The horizontal axis of the calibration curve represents the survival probabilities forecasted by the model. Conversely, the vertical axis illustrates the survival probabilities derived from the actual data. This configuration indicates that when the model’s predicted line at various time points aligns more closely with the gray line representing the ideal scenario, it signifies an improved predictive accuracy at those specific time points. The results showed that the clinical prognostic model of LUAD had the best clinical prediction performance for 3 years.

## 4. DISCUSSION

LUAD poses substantial threats to human health due to its aggressive nature and poor prognosis. The rising incidence and high mortality rate of LUAD underscore the urgent need for novel diagnostic and therapeutic strategies to improve patient outcomes [29, 30].

Investigating the phenotypic characteristics of LUAD and their underlying molecular mechanisms is crucial for advancing diagnostic and treatment approaches. Previous studies have shown that genetic and transcriptional profiling can reveal distinct properties of LUAD subtypes, aiding in the identification of potential biomarkers [31, 32]. The current research focuses on the differential expression of the PITPNC1 gene and its association with immune infiltration and clinical prognosis in LUAD. By leveraging comprehensive bioinformatics analyses and clinical data, this study aims to elucidate the potential of PITPNC1 as a key player in LUAD pathogenesis and its implications for personalized medicine [33, 34].

PITPNC1, a phosphatidylinositol transfer protein, is critically implicated in multiple cancer types, including LUAD. Recent research indicates that PITPNC1 is upregulated in LUAD and correlates with decreased patient survival. Mechanistically, PITPNC1 facilitates the interaction between KRAS and MYC, inhibiting autophagy through the mTOR pathway[35]. This regulatory function underscores a novel mechanism of KRAS-driven oncogenesis, presenting potential targets for therapeutic intervention. PITPNC1 enhances MYC protein stability and mTOR localization, crucial for tumor cell proliferation and survival. Targeting PITPNC1 could be a promising LUAD treatment strategy, potentially improving patient outcomes by disrupting key oncogenic pathways.

The study of the Hedgehog signaling pathway has revealed its critical role in development and tissue homeostasis, with its dysregulation being linked to various cancers and other diseases. The Hh pathway is essential for cell fate determination, proliferation, and differentiation, and its components include key molecules such as Smoothened (Smo) and Gli1, which are crucial for signal transduction. Aberrant Hh signaling has been implicated in the pathogenesis of liver fibrosis, where it modulates hepatic stellate cell (HSC) activation and extracellular matrix production, leading to fibrogenesis [36].

In the context of LUAD, the Hh pathway’s involvement in promoting cell proliferation and survival suggests that its dysregulation could contribute to tumorigenesis. The pathway’s role in maintaining stem cell populations and its interaction with other signaling mechanisms, such as Notch and IL12, underscores its potential as a therapeutic target. For instance, targeting the Hh pathway in combination with other treatments could enhance the efficacy of cancer therapies by disrupting the supportive tumor microenvironment and inhibiting cancer stem cell maintenance [37].

Furthermore, the Notch signaling pathway, another critical regulator of cell fate and differentiation, has been shown to interact with the Hh pathway. Notch signaling is involved in cell proliferation, apoptosis, and angiogenesis, and its dysregulation is associated with several cancers, including LUAD. The cross-talk between Notch and Hh signaling pathways can modulate the tumor microenvironment and influence cancer progression, highlighting the importance of understanding these interactions for developing effective therapeutic strategies [38].

In conclusion, the intricate interplay between the Hh, IL12, and Notch signaling pathways plays a significant role in the pathogenesis of LUAD. Targeting these pathways could provide new avenues for therapeutic intervention, potentially improving clinical outcomes for patients with LUAD. Further research into the molecular mechanisms underlying these interactions will be crucial for developing targeted therapies that can effectively disrupt tumor growth and progression.

The immune landscape of LUAD is complex and involves various immune cells that play crucial roles in tumor progression and patient prognosis. In our study, we observed significant differences in the infiltration of several immune cells, including eosinophils, across high and low PITPNC1 expression groups. Eosinophils, traditionally known for their role in allergic reactions and parasitic infections, have been increasingly recognized for their involvement in cancer. Specifically, eosinophilia has been associated with both tumor-promoting and tumor-suppressing activities. For instance, a study by Wang et al. highlighted that eosinophilia could potentially increase the risk of squamous cell lung cancer, suggesting a complex interplay between eosinophils and lung cancer pathogenesis[39]. Furthermore, eosinophil peroxidase (EPO), an enzyme abundantly expressed in eosinophils, has been shown to correlate with worse clinical outcomes in LUAD patients, indicating its potential as a prognostic marker[40].

The presence of eosinophils in the tumor microenvironment (TME) can influence tumor behavior and patient outcomes. Eosinophils contribute to the TME by releasing cytotoxic granules, cytokines, and chemokines, which can modulate immune responses and affect tumor growth. For example, eosinophil infiltration has been linked to poorer survival rates in LUAD patients, as demonstrated by Ye et al., where high EPO expression was associated with advanced tumor stages and lymph node metastasis[40]. This suggests that eosinophils may facilitate tumor progression through their interactions within the TME.

Our findings also indicated significant differences in the infiltration of other immune cells, such as CD56dim natural killer (NK) cells, which are known for their cytotoxic activity against tumor cells. Although no relevant literature was found directly linking CD56dim NK cells to LUAD, their role in immune surveillance and tumor eradication is well-documented in other cancers. The differential abundance of these immune cells in high and low PITPNC1 expression groups underscores the potential impact of PITPNC1 on the immune landscape of LUAD.

the varying infiltration patterns of eosinophils and other immune cells in LUAD highlights the intricate relationship between the immune system and tumor biology. The elevated presence of eosinophils in high PITPNC1 expression groups may contribute to a more immunosuppressive TME, promoting tumor progression and impacting patient prognosis. Understanding these interactions provides valuable insights into the potential mechanisms by which PITPNC1 influences LUAD and underscores the importance of considering immune cell infiltration in the development of targeted therapies.

## Conclusion

In summary, this study systematically elucidates the potential mechanisms and clinical significance of PITPNC1 in LUAD. Through various bioinformatics approaches, we confirmed the high expression of PITPNC1 in LUAD and demonstrated its potential as a diagnostic biomarker. Furthermore, we explored its association with immune infiltration, co-expressed genes, and clinical prognosis, providing a solid foundation for future research. These findings highlight the importance of PITPNC1 in LUAD and suggest that it could serve as a valuable target for diagnostic and therapeutic strategies in the future. Future studies should focus on validating these results through experimental and clinical approaches to fully realize the potential of PITPNC1 in LUAD management.

## Limitation

Despite the comprehensive bioinformatics analysis conducted in this study, several limitations should be acknowledged. Firstly, the study lacks validation through wet-lab experiments, which are crucial for confirming the bioinformatics predictions and understanding the underlying biological mechanisms. Secondly, although the sample size from TCGA and GEO databases is relatively large, it may still be insufficient to capture the full heterogeneity of LUAD. Thirdly, the absence of clinical validation, such as independent patient cohorts and prospective studies, limits the immediate clinical applicability of our findings. Additionally, the integration of multiple datasets could introduce batch effects despite the use of batch effect correction methods, potentially affecting the robustness of the results.

## Data Availability

All data produced in the present study are available upon reasonable request to the authors
All data produced in the present work are contained in the manuscript
All data produced are available online at

## AUTHOR CONTRIBUTIONS

Chao Li: conceived and designed the experiments and analyzed the data. JunsongChen and Ganggang Zhang: performed the experiments and prepared the figures and tables. Fang Guo: analyzed the data, revised it critically for important content. Xin Zhang: conceived and designed the experiments, analyzed the data, drafted the work.

## DATA AVAILABILITY STATEMENT

All raw data and code are available upon request.

## ETHICS STATEMENT

Not Applicable

